# Evaluation of ensilication technology for ambient DNA preservation

**DOI:** 10.1101/2025.10.30.25339175

**Authors:** Michael Blas, Celeste Yu, Farnoosh Abbas Aghababazadeh, Vanessa Hoelscher, Marcus Volz, Linden Morales, Kaytee Jarolik, Xiuhua Dong, Sheila Dobin, Tuula Rantasalo, Tatu Hirvonen, Juha-Pekka Pursiheimo, Nea Laine, Nils Homer, Cassandra Chamoun, William Pierce, Lee Organick, Adrian Fehr, Michael Becich, James L. Banal, Kenneth Youens, Manu Tamminen, Benjamin Haibe-Kains, Philippe Bedard

**Affiliations:** Princess Margaret Cancer Centre, University Health Network, Toronto, Ontario, M5G 2C1, Canada; Medical Biophysics, University of Toronto, Toronto, Ontario, M5G 2C4, Canada; Structural Genomics Consortium, Toronto, Ontario, M5G 1L7, Canada; Department of Computer Science, University of Toronto, Toronto, Canada; Vector Institute for Artificial Intelligence, Toronto, Ontario, M5G 0C6, Canada; Baylor Scott & White Health, Temple, TX, USA; Genomill Health Inc., Turku, 20520 Finland; Cache DNA, Inc. San Carlos, CA 94070 USA; Fulcrum Genomics LLC, Boston, MA 02144 USA

## Abstract

Current nucleic acid preservation relies on ultra-low temperature storage (−20 °C to −80 °C), imposing significant infrastructure, cost, and accessibility barriers that limit genomic medicine worldwide. We present a comprehensive evaluation of ensilication, a silica-based encapsulation method enabling ambient-temperature preservation of DNA without compromising sequencing fidelity. Across clinical, genomic, and biochemical analyses, ensilication maintained complete diagnostic concordance with cryogenic controls, detecting all actionable variants in FFPE tumor samples, even at low variant allele frequencies. Whole-genome sequencing revealed that frozen storage accumulated up to 65% more artifactual C>T mutations than ensilicated samples, underscoring its potential to reduce false-positive calls in oncology. Both linear and circular DNA libraries preserved structural integrity across temperatures from −80 °C to 37 °C. By eliminating cold-chain dependence, ensilication enables decentralized biobanking, point-of-care testing, and equitable access to precision oncology, enabling globally accessible cancer genomics. Its compatibility with emerging sequencing platforms positions ensilication as a foundational technology for next-generation diagnostics and large-scale population studies.

## Introduction

Current nucleic acid preservation methods require storage at temperatures that range from -20 °C to -140 °C to prevent hydrolytic and oxidative degradation. While cryogenic storage effectively arrests molecular motion and chemical reactivity, it requires continuous temperature maintenance and specialized infrastructure that limits biological research accessibility and geographic distribution. As genomic studies expand to include diverse populations and longitudinal sampling, the logistical constraints of cold chain dependence increasingly conflict with the need for distributed, large-scale biosample collection. Furthermore, temperature excursions during transport or equipment failure can compromise sample integrity^1–3^.

Several ambient-temperature preservation technologies have emerged as potential alternatives, including lyophilization, anhydrous storage matrices, and biomimetic encapsulation methods^4,5^. While these approaches demonstrate varying degrees of success in preventing nucleic acid degradation, existing evaluations primarily rely on indirect assessment methods such as gel electrophoresis^6^ for fragment sizing, qPCR for amplifiable template quantification^7,8^, or mass spectrometry for detecting specific chemical modifications^9^. When sequencing is employed, studies typically examine short synthetic oligonucleotides or plasmid DNA. While cost-effective, these model systems lack the complexity of genomic DNA with its repetitive elements, variable GC content, chromatin-associated modifications, and megabase-scale structural features.

Preservation artifacts that are negligible in a 100-base oligomer may become systematic biases when amplified across a 3-billion-base genome. For example, GC-rich regions might preferentially degrade^10–12^, repetitive sequences could cross-link^13–15^, or sequence-specific chemical modifications might accumulate in regulatory regions^10,16,17^. These concerns are substantiated by documented artifacts in existing preservation methods. Formalin fixation introduces C>T transitions through cytosine deamination that are routinely mistaken for somatic mutations^18–20^, while oxidative damage during desiccation generates G>T transversions indistinguishable from cancer driver mutations^21–23^. Even subtle preservation-induced biases in GC content or AT dropout could obscure clinically actionable variants or create false-positive mutation calls^24,25^.

Ensilication, which encapsulates nucleic acids within silica, may provide a chemically inert environment that preserves DNA without introducing chemical modifications^26^. However, whether this preservation method maintains the sequencing fidelity required for clinical diagnostics, particularly for challenging sample types like formalin-fixed paraffin-embedded (FFPE) tissues, remains unestablished. Here, we evaluate ensilication as an ambient-temperature storage method through comprehensive genomic analyses of matched clinical samples. We compare mutation detection, whole-genome quality metrics, and mutational signatures between ensilicated and frozen FFPE tumor samples, assess preservation efficacy across multiple temperature conditions, and examine both linear and circular DNA integrity. Our results demonstrate that ensilication enables ambient-temperature storage while maintaining complete diagnostic concordance with frozen controls, establishing it as a viable alternative to cryogenic preservation for clinical genomic applications.

## Results and Discussion

### Comparative performance of frozen and ensilicated storage using clinical mutation detection assays

Clinical genomics workflows demand absolute preservation of mutation detection capabilities, as even minor storage-induced artifacts can compromise therapeutic decision-making. To determine whether ensilication preserves clinically actionable mutation detection comparably to frozen storage, we analyzed ten FFPE-extracted DNA samples using the Agena iPLEX HS Lung Panel, a MALDI-TOF-based laboratory-developed test in routine clinical use at Baylor Scott & White Health for non-small cell lung cancer molecular diagnostics^27^. We selected archived specimens with known mutation status to challenge both storage methods across the dynamic range of clinical testing. The cohort included seven samples harboring driver mutations, namely RAS variants (G12C, G12V, Q61H), EGFR exon 19 deletions (L747_P753>S, E746_A750del, L747_S752del), and BRAF G469V, alongside three mutation-negative controls. This design encompassed both high-frequency alterations readily detected above 10% VAF and challenging low-frequency variants near detection thresholds (2–10% VAF), representing the full analytical spectrum encountered in clinical practice.

Following DNA extraction, matched aliquots were subjected to either standard cryogenic storage at -80 °C or ensilication at room temperature for 14 days. Post-storage analysis with technical replicates revealed complete diagnostic concordance between preservation methods. All seven mutation-positive samples retained their clinically actionable variants with identical calls, while mutation-negative controls generated no false positives under either condition. This 100% concordance extended even to variants at the lower detection range. KRAS Q61H hovering near 2% VAF and KRAS G12C at 4% were reliably detected regardless of storage method. Quantitative assessment of variant allele frequencies demonstrated remarkable preservation fidelity. Among the four samples with VAFs in the quantifiable 2-10% range, storage method introduced minimal variation: the mean absolute VAF difference was 0.42%, with the largest individual deviation reaching 1.12% for KRAS G12C. These results demonstrate that ensilication, despite its fundamentally different preservation chemistry from cryogenic storage, maintains the DNA integrity required for clinical mutation detection.

**Table 1.** Clinical mutation detection concordance between frozen and ensilicated storage methods. Ten FFPE-extracted DNA samples were analyzed using the Agena iPLEX HS Lung Panel with technical replicates for each storage condition. VAF, variant allele frequency. n.d., non-detected.

| Sample | Mutation | Frozen VAF replicate 1 [%] | Frozen VAF replicate 2 [%] | Ensilicated VAF replicate 1 [%] | Ensilicated VAF replicate 2 [%] |
| --- | --- | --- | --- | --- | --- |
| 1 | KRAS Q61H | 2.11 | 2.20 | <2 | 2.28 |
| 2 | EGFR L747_P753>S | >10 | >10 | >10 | >10 |
| 3 | No mutation | n.d. | n.d. | n.d. | n.d. |
| 4 | EGFR E746_A750del | >10 | >10 | >10 | >10 |
| 5 | EGFR L747_S752del | >10 | >10 | >10 | >10 |
| 6 | KRAS G12C | 4.30 | 4.56 | 3.18 | 4.27 |
| 7 | No mutation | n.d. | n.d. | n.d. | n.d. |
| 8 | KRAS G12V | 7.33 | 6.94 | 7.42 | 7.68 |
| 9 | No mutation | n.d. | n.d. | n.d. | n.d. |
| 10 | BRAF G469V | 8.71 | 8.39 | 8.47 | 8.59 |

### Storage-dependent mutation accumulation in FFPE-extracted DNA impacts clinical variant detection

While targeted clinical assays demonstrated equivalent performance, they interrogate only high-frequency variants at specific loci, potentially missing the accumulation of low-frequency mutations across the genome that may reveal storage-specific chemical degradation. As clinical genomics evolves toward comprehensive profiling, namely assessing tumor mutational burden, signature analysis, and subclonal architecture, understanding these genome-wide storage artifacts becomes essential. We therefore performed deep whole-genome sequencing (30× normal, 80× tumor) to characterize the full spectrum of storage-induced mutations invisible to targeted platforms.

FFPE blocks from 32 matched normal-tumor pairs were obtained from Ontario Tumor Bank. DNA was extracted, divided into aliquots, and stored either frozen at -80 °C or ensilicated at room temperature for 28 days before sequencing. Despite divergent storage chemistries, both methods preserved DNA integrity equally well, with virtually identical sequencing metrics including insert sizes, chimera rates, and library complexity across tissue types (**Supplementary Tables 1–3**). This technical equivalence enabled direct comparison of storage-induced mutational changes without confounding quality differences.

The mutational landscapes revealed storage-dependent patterns. While normal tissue DNA showed minimal changes under both conditions (**Figure 2a**, **Supplementary** Figures 1**, 3,** and **5**), tumor DNA exhibited different accumulation patterns. Frozen storage permitted C>T accumulation reaching 5,466 mutations per million bases, compared to 3,307 following ensilication, a 65% increase representing over 2,000 additional mutations (p=0.006, **Supplementary Table 5**). This represents over 2,000 additional artifactual mutations per million bases that would be indistinguishable from true somatic variants. Sequence context analysis revealed these C>T mutations concentrated at CpG dinucleotides, exceeding 4,000 mutations per million bases in NCG trinucleotide contexts (**Supplementary** Figures 2**, 4,** and **6**). These CpG-context C>T transitions are well-established sources of false-positive somatic mutation calls in clinical FFPE sequencing, particularly at recurrent cancer driver mutation sites. This pattern demonstrates that FFPE-initiated cytosine deamination continues even at -80 °C but is arrested by ensilication, suggesting fundamental differences in how these storage methods affect DNA chemistry.

**Figure 2.**
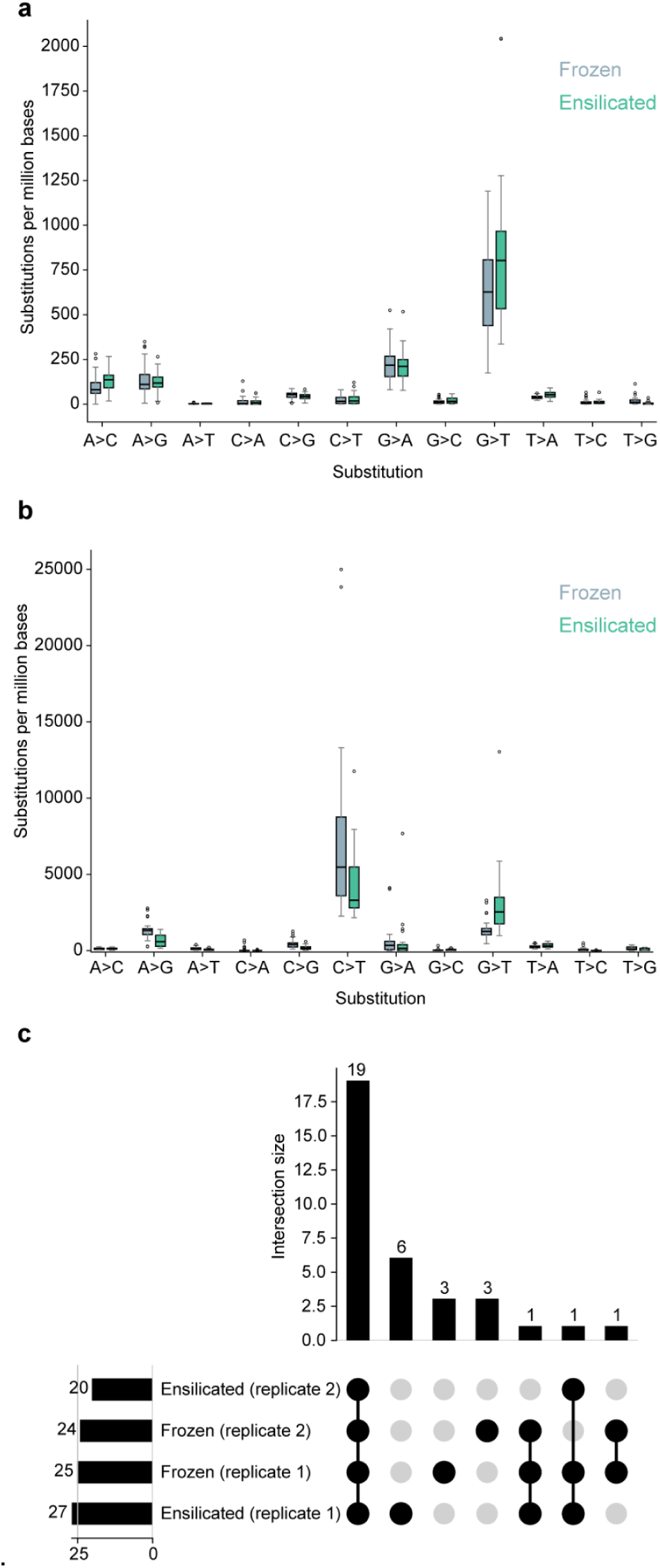
Storage-dependent mutation accumulation in FFPE-extracted DNA and concordance of clinical variant detection. a, Mutation frequencies across substitution types in normal tissue DNA following frozen (−80°C) or ensilicated (room temperature) storage. n = 32 for both storage conditions. **b,** Mutation frequencies in tumor tissue DNA under identical storage conditions, showing pronounced C>T accumulation in frozen samples and G>T elevation in ensilicated samples. n = 32 (frozen) and 29 (ensilicated). Box plots show median, interquartile range, and 1.5× IQR whiskers; outliers shown as individual points. **c,** UpSet plot showing concordance of 34 oncogenic mutation calls across technical replicates. Intersection size indicates number of mutations detected in specific replicate combinations. Connected dots indicate replicate membership for each intersection.

Clinical variant detection remained robust under both storage conditions. Among 34 evaluated oncogenic mutations, concordance between replicates was high, with discrepancies primarily affecting low-frequency variants (**Figure 2c**). Each storage method showed single instances of replicate dropout at intermediate frequencies (10% for ensilication, 20% for frozen), while variants below 5% showed expected stochastic variation. The notable exception was KRAS G12D, detected at 20% allele frequency in both frozen replicates but absent from ensilicated samples. This C>T transition notably matches the predominant mutation type accumulating in frozen storage. The overall high concordance indicates that while storage profoundly affects background mutation rates, clinical variant calling remains largely preserved.

### Ensilication preserves DNA integrity regardless of topology

Conventional DNA libraries are composed of linear double-stranded DNA molecules, which inherently suffer from limited stability and suboptimal long-term preservation^5,28^. Recently, several next-generation sequencing technologies, such as Pacific Biosciences Onso^29^, Element Biosciences AVITI^30^, and MGI DNBSEQ^31^ sequencing platforms, have introduced workflows that use circular ssDNA (cssDNA) intermediates. Moreover, circular DNA library intermediates confer clear benefits for nanopore sequencing, particularly by enhancing structural stability and enabling continuous long-read generation^32^.

To address these developments, we established a novel method for generating cssDNA libraries that are directly compatible with the aforementioned sequencing platforms^33^. Owing to their covalently closed topology, circular DNA molecules lack free reactive 3’ and 5’ ends and are also less susceptible to nuclease activity and spontaneous degradation^5^. Building on this inherent structural stability, we evaluated whether these cssDNA molecules could be ensilicated and compared their stability under various storage conditions relative to linear DNA molecules. This study aimed to determine whether cssDNA libraries provide superior preservation and functional integrity during long-term storage and subsequent handling.

To investigate whether ssDNA libraries can be encapsulated and whether their molecular conformation influences the preservation, we analyzed the stability of linear ssDNA and cssDNA molecules subjected to different storage conditions. Both library types were incubated under various storage settings for 20 hours. The integrity of the DNA molecules was evaluated by generating Illumina-compatible sequencing libraries through PCR, followed by analysis of PCR products using agarose gel electrophoresis. The resulting libraries were further assessed by sequencing on an Illumina MiSeq platform to determine potential structural or functionality-related differences between the linear and circular forms and between different storage conditions.

Agarose gel electrophoresis revealed that the ensilication efficiently preserved both linear and cssDNA molecules across all tested conditions (**Supplementary Table 6**). Ensilicated samples showed no detectable degradation, confirming that the matrix provides a robust physical and chemical barrier against DNA hydrolysis and oxidation. Notably, cssDNA exhibited remarkable inherent stability even in the absence of encapsulation, showing minimal or no visible degradation across all tested conditions. In contrast, for linear ssDNA, encapsulation provided a clear advantage, offering strong protection and markedly outperforming unprotected samples under every temperature regime, specifically -80 °C, -20 °C, ambient temperature, and 37 °C, throughout the 20-day incubation period. These results highlight the ability of the ensilication process to maintain the structural integrity of even the more degradation-prone linear DNA species, while also emphasizing the intrinsic resilience of circular DNA molecules.

To accurately evaluate the impact of storage conditions on DNA integrity, sequencing libraries were analyzed on an Illumina MiSeq platform. Sequencing data revealed that, after normalization of input concentrations, libraries generated from cssDNA consistently yielded higher total read counts than those derived from linear DNA (**Figure 3a**). This observation indicates that a larger fraction of cssDNA molecules retained their amplifiable and sequenceable structure after storage. Furthermore, read depth variance across samples closely correlated with the relative band intensities observed in agarose gels, validating the visual gel-based observations through sequencing-based quantification (**Supplementary Table 6**).

**Figure 3.**
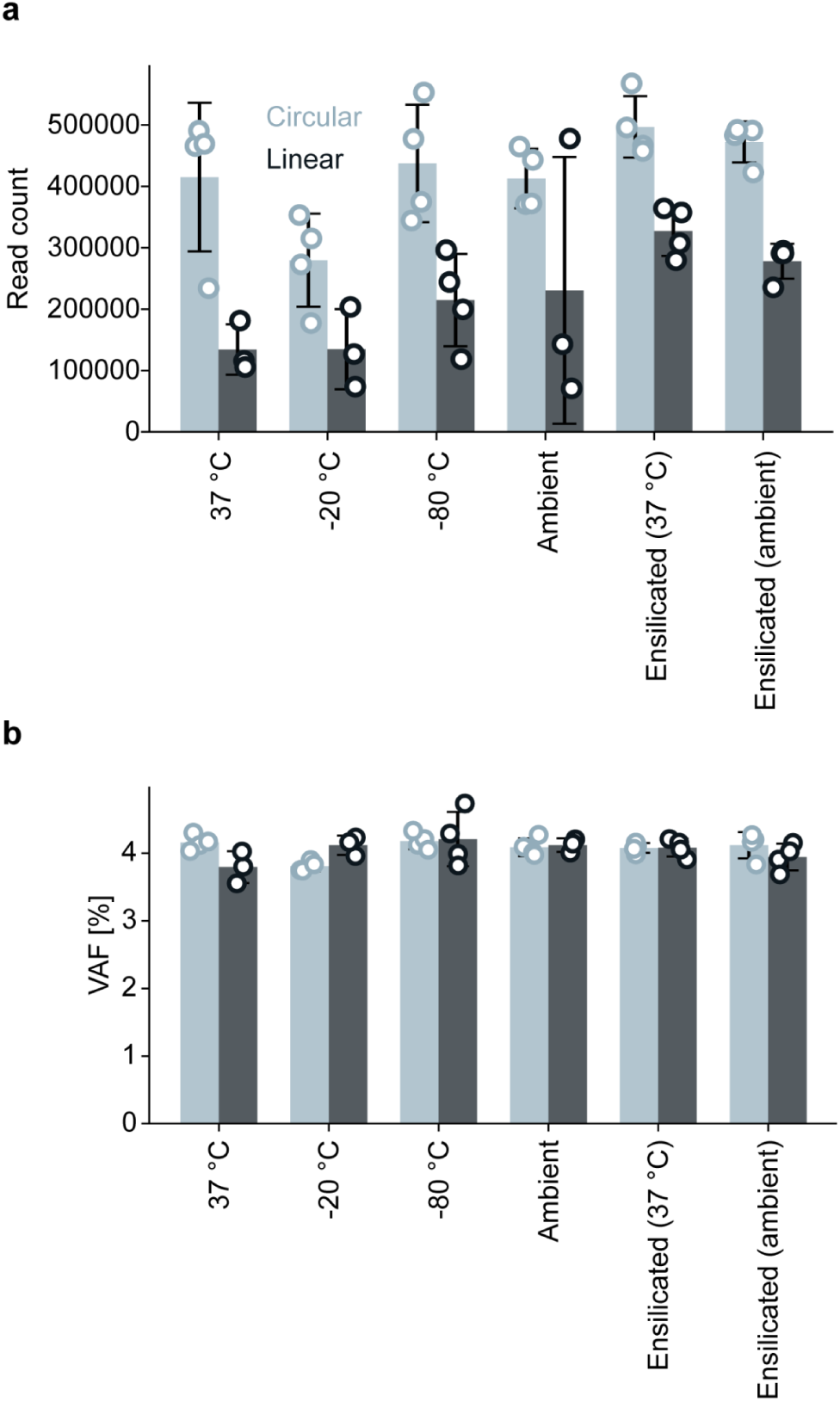
Comparison of DNA stability across storage methods and conditions. a, Sequencing read depths. Error bars show standard deviation between 4 replicates. **b,** Observed variant allele frequencies (VAF) (%). Error bars show standard deviation between 4 replicates. Bar height denotes average.

Variant analysis provided additional insights into sequence fidelity. Across all storage conditions and molecular topologies, variants present at 5% frequencies were accurately detected (**Figure 3b**). This finding demonstrates that the sequence composition of both library types remained intact, even in samples that experienced partial loss of amplifiable molecules. Thus, while total yield and functional integrity may vary, the base-level sequence integrity of surviving DNA molecules appears unaffected by either storage condition or DNA topology. Nevertheless, a clear reduction in the number of functional molecules was observed in linear DNA libraries compared to circular ones, emphasizing that linear DNA is substantially more susceptible to damage during storage. The observed losses likely result from strand scission and depurination events initiated at free DNA ends, which are mechanisms inherently mitigated in circular topologies. These findings collectively support the conclusion that cssDNA molecules possess intrinsic structural advantages that confer enhanced protection during both ensilicated and non-ensilicated storage.

### Broader considerations towards implementing ambient nucleic acid storage

Today, sample storage relies heavily on a distributed infrastructure of -20 °C and -80 °C freezers, complemented by liquid handlers, consumables (pipette tips, 96-well transfer plates, boxes, and racks)–which scale linearly with throughput—and significant technician labor. Each freezer carries not only its purchase and amortization costs but also recurring expenses such as electricity, preventive maintenance contracts, temperature monitoring systems, insurance, and real estate requirements. While the initial purchase price of an ultra-low temperature (ULT) freezer can range from $7,000-$35,000 USD^34,35^, this represents only 28% of the lifetime cost (39% is energy consumption, 11% is maintenance, 11% is HVAC, and 11% is floor space)^36,37^.

The National Institutes of Health (NIH), which has over 3,400 ULT freezers in service, points out that the environmental impact of these units amounts to 18,000 lbs. of CO_2_ per year^38^.

Beyond performance characteristics between ensilication and cold storage approaches, several other factors remain barriers to the implementation of ambient nucleic acid preservation. A total cost of ownership (TCO) analysis must account for the entire lifecycle of biospecimen management—from sample accession to long-term archival—and quantify both direct and indirect costs associated with ambient and current cold-chain workflows. Indirect costs such as HVAC load, emergency power supply, and compliance-related monitoring further inflate the overall expense but are often hidden in overhead budgets. Uncertainty of grant funding and long time horizons from sample collection to utilization have led many groups, such as the National Cancer Institute and University of British Columbia (UBC), to create tools for estimating the economics of biobanking and associated laboratory operations^39,40^

While ensilication (as described) shows promise in preserving DNA integrity and sequence fidelity, the additional complexity introduced to laboratory operations by an extended shaking step (up to 2 days) is not conducive to most sample handling workflows. Furthermore, materials safety risks with the reagents necessary for the retrieval of samples are suboptimal for wide distribution. Not only must a solution demonstrate robust stability, sufficient recovery, and sequencing concordance data to be adopted, but methods must be of similar or lower complexity and duration to cold storage equivalents, be able to be delivered in manual and automated formats, and yield a compelling return-on-investment to laboratory operators considering ambient storage for prospective and retrospective samples.

Limited data exists examining the utility and cost-benefit analysis of transitioning from cold storage to room temperature alternatives. Future studies, capturing real-world data and benchmarking against the performance of ensilication, are necessary to evaluate chemical formulations that can enable broader implementation of ambient nucleic acid storage.

### Conclusion and Outlook

This study provides a comprehensive genomic evaluation of ensilication using direct sequencing rather than indirect preservation metrics. The Agena iPLEX HS Lung Panel demonstrated complete diagnostic concordance, detecting all seven clinically actionable mutations in both ensilicated and frozen FFPE samples after 14 days, including low-frequency variants at 2–10% VAF. Whole-genome sequencing of 32 tumor-normal pairs quantified preservation-induced changes. Frozen samples at -80 °C showed accumulation of C>T mutations after 28 days of storage, while ensilicated samples at room temperature showed fewer. Further, ensilicated samples showed concordance in detecting oncogenic mutations. Evaluation of DNA topology revealed that ensilication preserved both linear and circular single-stranded DNA across all tested temperatures (-80 °C to 37 °C) over 20 days, with linear DNA showing marked improvement with encapsulation while circular DNA maintained integrity even unprotected, and all samples accurately detected 5% frequency variants regardless of storage condition or DNA topology.

These findings establish ensilication as a potential path toward decentralizing genomic medicine. Room-temperature preservation that maintains clinical diagnostic quality removes the primary infrastructure barrier preventing genomic testing in remote locations, developing nations, and point-of-care settings. Ensilication’s compatibility with both traditional and emerging sequencing library formats positions it for current clinical workflows and next-generation platforms. Realizing this potential requires extending validation across temporal and biological dimensions. Preliminary longitudinal studies show stable DNA integrity scores through 15 weeks at room temperature (**Supplementary** Figure 7). Multi-year stability studies will establish suitability for biobanking applications and define optimal storage windows. Validation across tissue types, extraction methods, and DNA quality grades will demonstrate universal applicability of ensilication. Protocol optimization for sample input, encapsulation conditions, and recovery procedures will streamline clinical integration. As these studies progress, early adoption in sample transport applications, where the 28-day stability window already exceeds typical shipping timeframes, can demonstrate real-world performance while building confidence for broader implementation. The convergence of ambient-temperature preservation with advancing sequencing technologies will ultimately enable genomic medicine to reach any location where samples can be collected.

## Methods

### Clinical assay sample processing and storage

Ten genomic DNA samples were analyzed upon receipt using 10 μl quality control aliquots. Initial characterization included Qubit dsDNA High Sensitivity assay for concentration determination, TapeStation 4200 for DNA integrity assessment (DIN scores), and NanoDrop for purity evaluation (260/280 and 260/230 ratios). Based on measured concentrations, volumes containing 1 μg of DNA were calculated for each sample.

For ensilication, 1 μg of DNA from each sample was subjected to the ensilication reaction with orbital shaking for 48 h. Following shaking, ensilicated DNA was pelleted by centrifugation, supernatant was removed, and pellets were resuspended in fresh storage buffer. Samples were then maintained at ambient temperature (22-25°C) for 14 days. Parallel control aliquots containing 1 μg DNA were prepared simultaneously and stored at -80 °C for the study duration.

After 14 days of storage, ensilicated DNA was recovered by pelleting samples, removing storage buffer, and adding oxide etch buffer. Samples were incubated at 40°C for 10 min to ensure complete dissolution of the silica matrix. Complete silica removal was verified by visual inspection following centrifugation. Released DNA was purified using Amicon Ultra 0.5 ml centrifugal filters (30 kDa MWCO) to remove residual impurities. Post-recovery quality control included Qubit quantification, TapeStation integrity analysis, and NanoDrop purity assessment. Sample volumes were measured to calculate total DNA mass recovery. Frozen control samples were thawed and analyzed using identical quality control metrics. Both filtered ensilicated samples and thawed frozen controls were divided into equal-volume aliquots for downstream analysis.

### Whole genome sequencing sample processing and storage

Sixteen matched patient sample sets, each comprising normal and tumor DNA, were processed for storage comparison studies. Upon receipt, 10 μl quality control aliquots were prepared from each sample. DNA samples were characterized using Qubit dsDNA High Sensitivity assay for quantification, Genomic DNA TapeStation for integrity assessment (DIN scores), and NanoDrop for purity evaluation. Each sample was divided equally by volume for parallel processing.

Half of each sample volume underwent ensilication with orbital shaking for 48 h. Following pelleting by centrifugation and supernatant removal, ensilicated samples were resuspended in fresh storage buffer and maintained at ambient temperature (22-25°C) for 28 days. The remaining half of each sample was stored at -80 °C in original tubes as controls.

After 28 days, ensilicated samples were recovered using the same pelleting, buffer exchange, and oxide etch protocol described above (40°C, 10 min). Released nucleic acids were purified using Amicon Ultra centrifugal filters (30 kDa MWCO). For DNA samples, post-recovery quality was assessed using the same Qubit, TapeStation, and NanoDrop assays. Sample volumes were measured to calculate total mass recovery. Frozen control samples were thawed and analyzed using the same quality control metrics. Both ensilicated and control samples were aliquoted equally by volume for downstream sequencing analysis, with normal DNA sequenced at 30× coverage and tumor DNA at 80× coverage for whole genome sequencing.

### Whole genome sequencing data processing and alignment

Raw sequence data in FASTQ format were demultiplexed using Illumina’s bcl2fastq software bcl2fastq workflow (https://support.illumina.com/sequencing/sequencing_software/bcl2fastq-conversion-software.html). Per-sample FASTQs were trimmed with cutadapt (v.1.8.3)^41^ to remove adapter sequences, then aligned to the hg38 human reference genome (patch 12) with BWA-MEM (v.0.7.17)^42^.

Alignments were sorted, indexed, and marked for duplicates using samtools (v.1.9)^43^ and Picard MarkDuplicates (GATK v.4.1.6.0)^44^. Base quality scores were recalibrated with GATK BaseRecalibrator (GATK v.4.1.6.0)^44^. These tools were orchestrated using OICR’s Genome Sequence Informatics (GSI) bcl2fastq (https://github.com/oicr-gsi/bcl2fastq), bwaMem (https://github.com/oicr-gsi/bwaMem), and bwaMergeProcessing (https://github.com/oicr-gsi/bamMergePreprocessing) workflows.

### Whole genome sequencing variant calling and filtering

Variants were called using MuTect2 and FilterMutectCalls (GATK v.4.1.6.0)^44^ for variant detection and filtering. Functional annotation was performed with VEP (version 105.0)^45^, and converted to MAF with vcf2maf (version 1.6.21b) (https://github.com/mskcc/vcf2maf).These tools were orchestrated using OICR’s GSI mutect and variantEffectPredictor workflows.

### Whole genome sequencing library quality metrics

Library insert size, chimera rate, and complexity (duplication rate) were assessed on a per-sample basis using Picard (v.3.4.1)^44^ tools: CollectInsertSizeMetrics, CollectAlignmentSummaryMetrics, and MarkDuplicates, respectively (**Supplementary Tables 1–3**).

### Whole genome sequencing substitution rate analysis

For computational efficiency, reads aligned to chromosome 22 were extracted using samtools (v.1.2.1)^43^, on which Picard (v.3.4.1)^44^ CollectSequencingArtifactMetrics was run to compute error metrics. The detailed pre-adapter metrics from this tool were used to measure artifactual substitution rates that occur prior to adapter attachment during the PCR amplification process, e.g., shearing. Read counts for both the substitution (PRE_ALT_BASES) column and the refuting (CON_ALT_BASES) were collated via the overall error rate (ERROR_RATE), normalized to substitutions per million bases sequenced. These rates are presented in **Supplementary Tables 4–5** and **Supplementary** Figures 1–6.

### Converting sample DNA into Nicking Loop™ circular ssDNA library

A sample oligonucleotide pool containing 2.5 fmol of each target was mixed with 10 fmol of each probe and 200 fmol of Loop oligonucleotide in 1× Ampligase buffer (LGC Biosearch Technologies, Hoddesdon, UK). The mixture was denatured at 95 °C for 5 min and gradually cooled (10 °C decrease every 2.5 min) to 55°C to allow probe–target hybridization, followed by a 2 h incubation. Circularization was performed by adding 0.2 U Phusion High-Fidelity DNA Polymerase (Thermo Fisher Scientific, Waltham, MA) and 0.2 U Ampligase (LGC Biosearch Technologies) with 10 mM dNTPs (Thermo Fisher Scientific) in 1× Ampligase Reaction Buffer, and incubating at 55 °C for 40 min. Residual linear DNA was digested using 1 µl each of Thermolabile Exonuclease I, RecJf, and Lambda Exonuclease (NEB, Ipswich, MA) at 37 °C for 1 h, followed by enzyme inactivation at 80 °C for 10 min. The circular DNA library was purified with AMPure XP beads (Beckman Coulter, Brea, CA) using 2× bead volume and eluted in 35 µl of 55 °C nuclease-free water.

### Amplifying the circular ssDNA library and production of circular and linear libraries

To amplify the circularized molecules, 10 µl of Nicking Loop™-converted cssDNA (n = 20) was pre-annealed with 5 pmol of universal amplification primer in 1× EquiPhi29 buffer (Thermo Fisher Scientific). The mixture was incubated at 95°C for 3 minutes and then gradually cooled to room temperature to facilitate primer annealing to the cssDNA template. RCA was initiated by adding 10 mM dNTPs, 100 mM DTT and 0.5 U of EquiPhi29 (Thermo Fisher Scientific) followed by incubation at 45°C for 90 minutes. The reaction was terminated by heat inactivation of the enzyme at 95°C for 10 minutes. The newly synthetized concatemeric ssDNA was supplemented with nuclease-free water and 1× rCutSmart buffer (NEB). The supplemented reactions were heated to 95°C for 5 minutes and then gradually cooled to room temperature to promote Loop folding. The double-stranded segment of the folded Loops contained recognition site for nicking enzyme. The recognition site was digested with 10 U of Nb.BsrDI (NEB) at 37 °C for 90 minutes resulting into linear monomers. The enzyme was subsequently heat-inactivated at 80°C for 10 minutes. The resulting linear monomers were purified using 1.8× volume of AMPure XP beads (Beckman Coulter).

The purified linear ssDNA monomers were re-cirularized by diluting 10 µl of the monomer solution with 7 µl of nuclease-free water. To promote the self-annealing, the dilution was denaturated at 95°C for 5 minutes, after which the heating was turned off, letting the reaction to gradually cool down in 5 minutes, followed by cooling down in RT for 15 minutes. After denaturation the dilution was supplemented with 1× T4 ligase buffer and 1 µl of T4 DNA ligase (NEB) at 26 °C for 8 hours to ligate the self-annealed ssDNA monomers into complete circular ssDNA molecules. The ligation reaction was cleaned of residual ssDNA by adding 1 µl of Thermostabily Exonuclease I and RecJf (NEB) incubating the reaction at 37 °C for 45 min and terminating the reaction by heat inactivation after at 95°C for 10 minutes. The amplified circular library was purified with AMPure XP beads (Beckman Couter, Brea, CA) using 2× volume of beads and elution volume of 30 µl nuclease-free water.

To initiate re-circularization, 10 µl of linear monomers were mixed with 7 µl of nuclease-free water. The mixture was denatured at 95°C for 5 minutes and gradually cooled to room temperature to allow folding of Loop. Subsequently, 1× T4 DNA ligase buffer and 1 µl of T4 DNA ligase (NEB) were added to each reaction to ligate the nick in the folded monomer. Ligation was carried out at 32°C for 30 minutes. After the ligation, any residual ssDNA was removed by adding 1 µl each of Thermolabile Exonuclease I and RecJf (NEB). The reactions were incubated at 37 °C for 45 minutes, then heat-inactivated at 95°C for 10 minutes. The reactions were purified using 2× volume of AMPure XP beads (Beckman Coulter). After this step, the original cssDNA molecules had been successfully amplified resulting in a first-generation of amplified molecules. This entire process—RCA, loop digestion, and re-circularization—was repeated up to three times using the newly generated circular molecules as templates and a reverse amplification primer to produce subsequent generations of cssDNA molecules.

In the course of the third amplification round, a subsample of purified linear ssDNA monomers was collected at the intermediate stage of the reaction to serve as the linear library for storage experiments. The remaining reaction was continued to completion, yielding the circular library used in parallel. To ensure comparability between samples, all libraries were stored at −80 °C prior to the start of the storage experiment.

### Storage conditions

Linear and circular DNA libraries were transferred into storage buffer (10 mM Tris (pH 7.5), 1 mM EDTA, and 50 mM NaCl). To minimize variability from repeated freeze–thaw cycles, all libraries were thawed simultaneously at the start of the storage study. The libraries awaiting the processing of parallel libraries were kept at 4 °C for up to two days before transfer to their designated storage conditions. All libraries were stored in 1.5 mL Eppendorf snap-cap tubes sealed with parafilm to reduce evaporation.

### Temperature-based storage of linear and circular DNA libraries

Libraries were stored at –80°C, –20°C, ambient temperature or 37 °C for 20 days. No additional treatments were applied prior to storage. Upon completion of the storage period, Illumina sequencing libraries were prepared directly from samples corresponding to each storage condition.

### Ensilication storage of linear and circular DNA libraries

For libraries stored at ambient temperature or 37 °C with encapsulation, the entire library volume was processed using the Cache kit according to the manufacturer’s protocol. De-encapsulation was carried out following the manufacturer’s instructions with two modifications: (i) in DCAP Reaction step 3c, the volume of the etching solution was increased to 400 µl and (ii) in the Buffer Exchange protocol step 1, the volumes were adjusted to 100 µl of Diluent B and 400 µl of de-encapsulated sample.

Following de-encapsulation, the libraries were immediately used for Illumina library preparation.

### Illumina library preparation and sequencing of linear and circular DNA

After the storage period, 2µl of each library (linear or circular) was used to prepare Illumina sequencing libraries. Amplification was carried out by PCR with Phusion HS II DNA Polymerase (Thermo Fisher Scientific) and 10 μM dual-indexed primers. The thermal cycling program consisted of 15 cycles including an annealing step at 68 °C.

PCR products were separated by gel electrophoresis, and target fragments were purified using the Monarch DNA Gel Extraction Kit (NEB). Purified libraries were quantified using the Qubit™ dsDNA HS Assay Kit and measured with a Qubit™ 4 Fluorometer (Thermo Fisher Scientific).

Sequencing was carried out on the Illumina MiSeq platform (MiSeq Reagent Kit v3, 2 × 300 bp). Three libraries were excluded from the run due to low quality: one replicate each from 3 7°C linear, -20 °C linear and ambient linear conditions.

### Sequencing data analysis of linear and circular DNA libraries

Reads were merged using VSEARCH (v2.15.2_linux_x86_64)^1^ with the following parameters: --fastq_minovlen 10 --fastq_maxdiffs 15 --fastq_maxee 1 --fastq_allowmergestagger --fastq_qmaxout 92. Otherwise, data were processed using Genomill’s proprietary pipeline. For **Figure 3**, python version 3.11.5 and matplotlib library version 3.7.2 were used.

### Ethics Declaration Statement

Baylor Scott & White Health ethical approval was obtained from the Baylor Scott & White Health Institutional Review Board (protocol 024-276). Ethical and research approvals for participants in Ontario-wide Cancer TArgeted Nucleic Acid Evaluation (OCTANE) were obtained from the Ontario Cancer Research Ethics Board (project ID 1217). All participants provided written informed consent.

## Acknowledgements

The Ontario-wide Cancer TArgeted Nucleic Acid Evaluation (OCTANE) study was conducted with the support of the Ontario Institute for Cancer Research through funding provided by the Government of Ontario (grant numbers P.OCT.051 and P.AO.075) and by the Princess Margaret Cancer Foundation. Baylor Scott & White Health biorepository resources were provided in part by a grant from the Baylor Scott & White Central Texas Foundation. Part of the work was financially supported by the Li Ka Shing Foundation.

## Code availability

The code used for data analysis is available at 10.5281/zenodo.17469053.

## Data availability

Data will be submitted to the European Genome-phenome Archive (EGA) upon publication.

## Supplementary Information

**Supplementary Figure 1.**
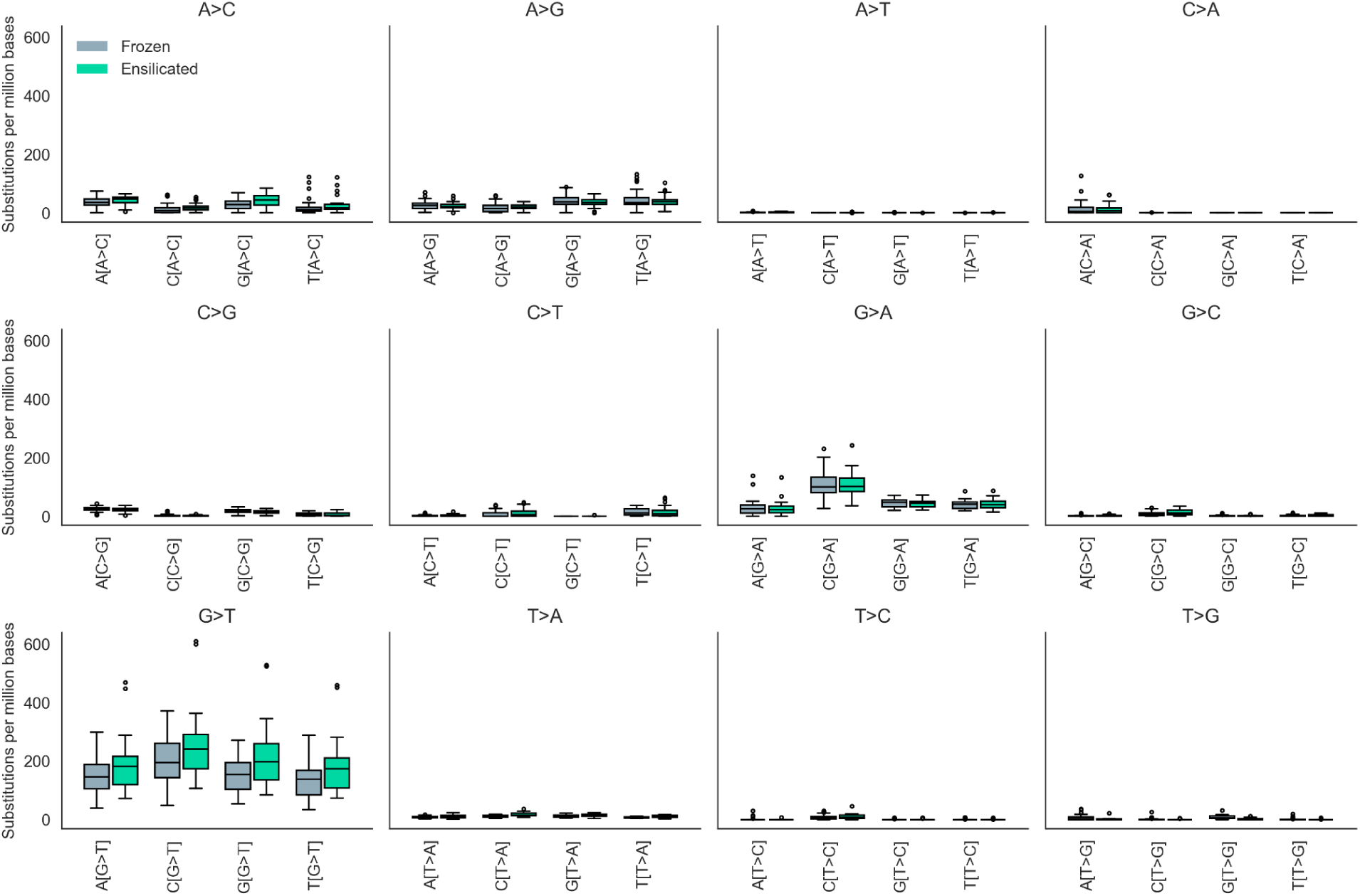
Substitution rates with pre-dinucleotide context from normal tissue.

**Supplementary Figure 2.**
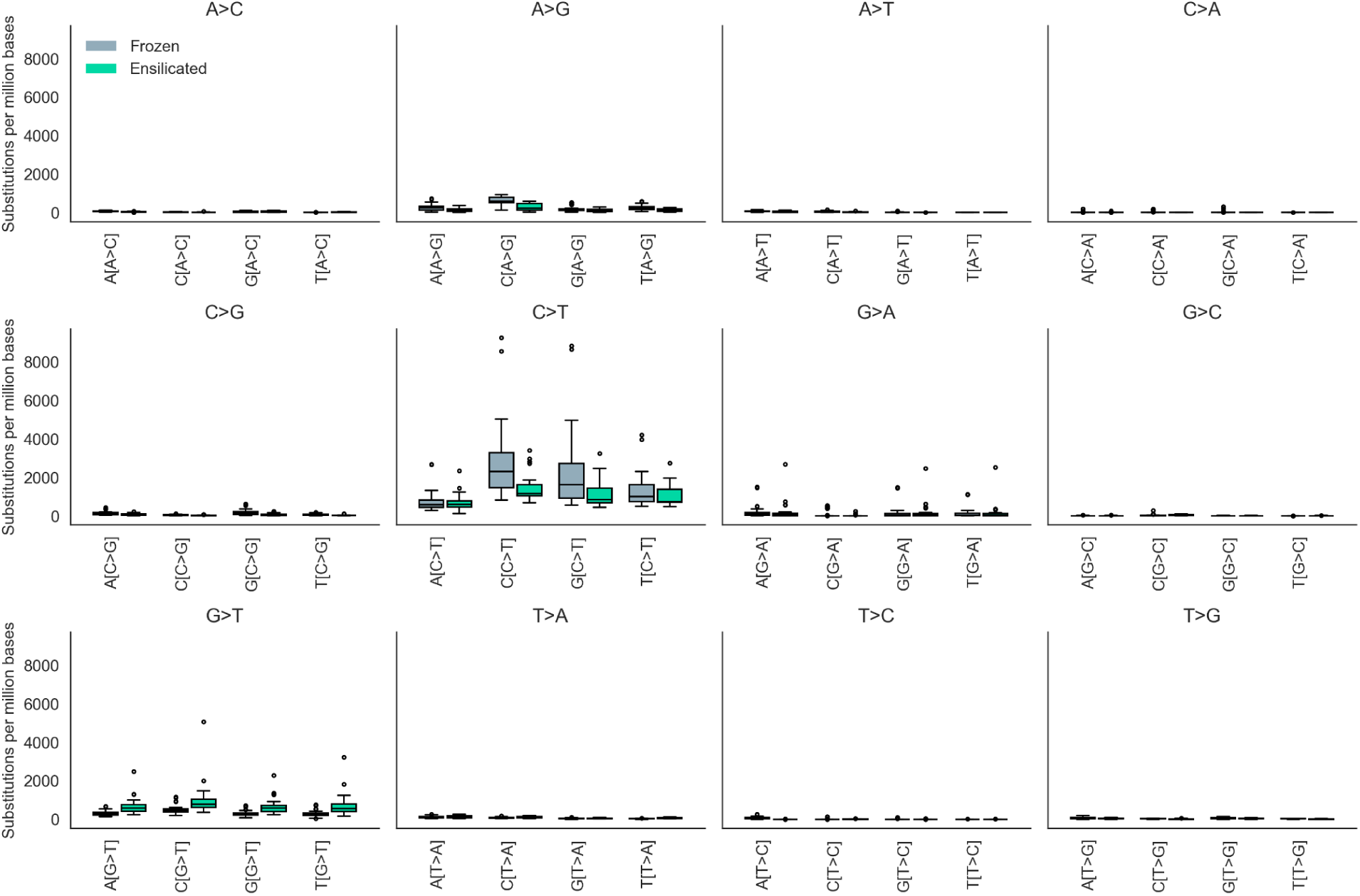
Substitution rates with pre-dinucleotide context from tumor tissue.

**Supplementary Figure 3.**
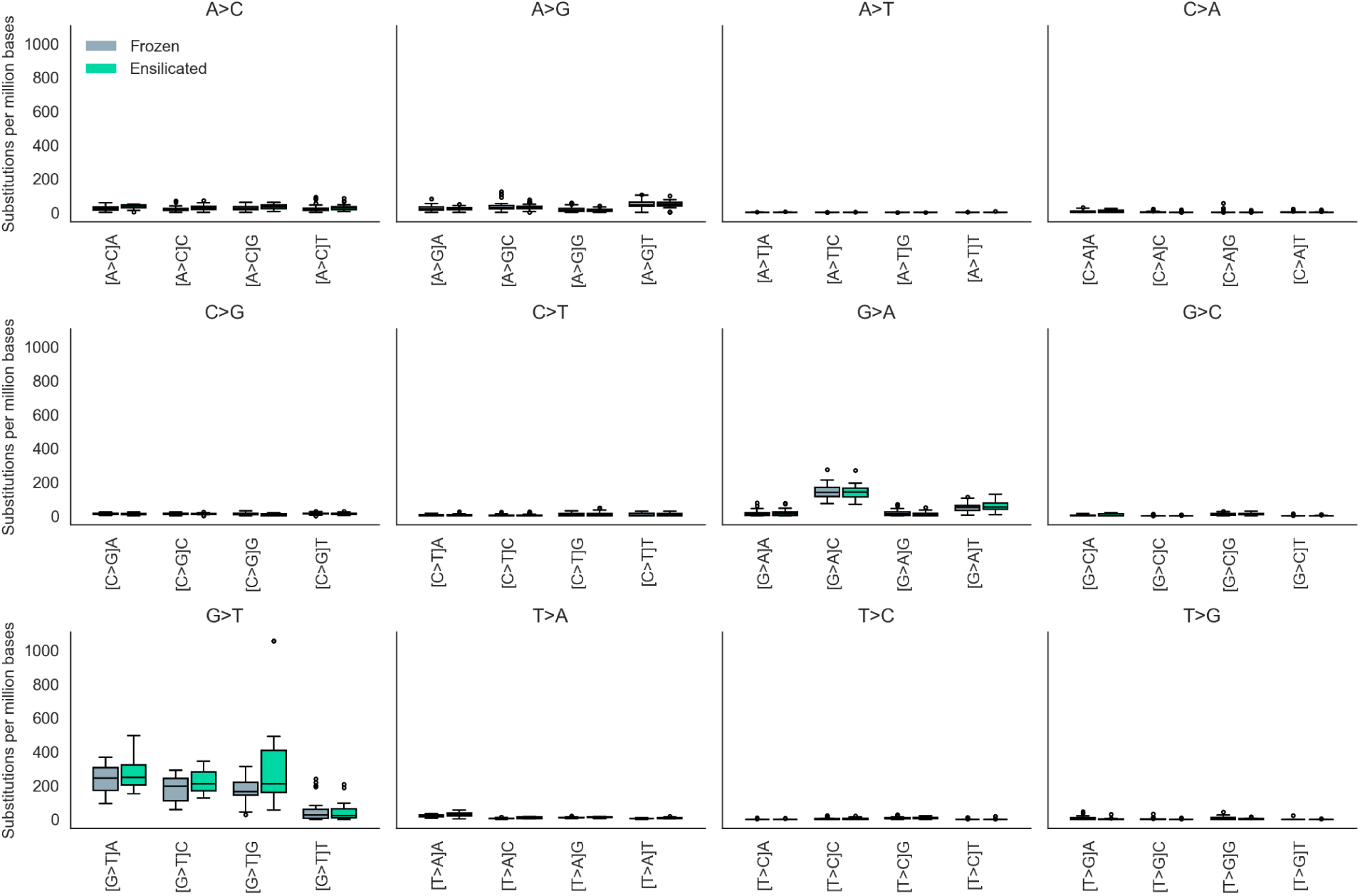
Substitution rates with post-dinucleotide context from normal tissue.

**Supplementary Figure 4.**
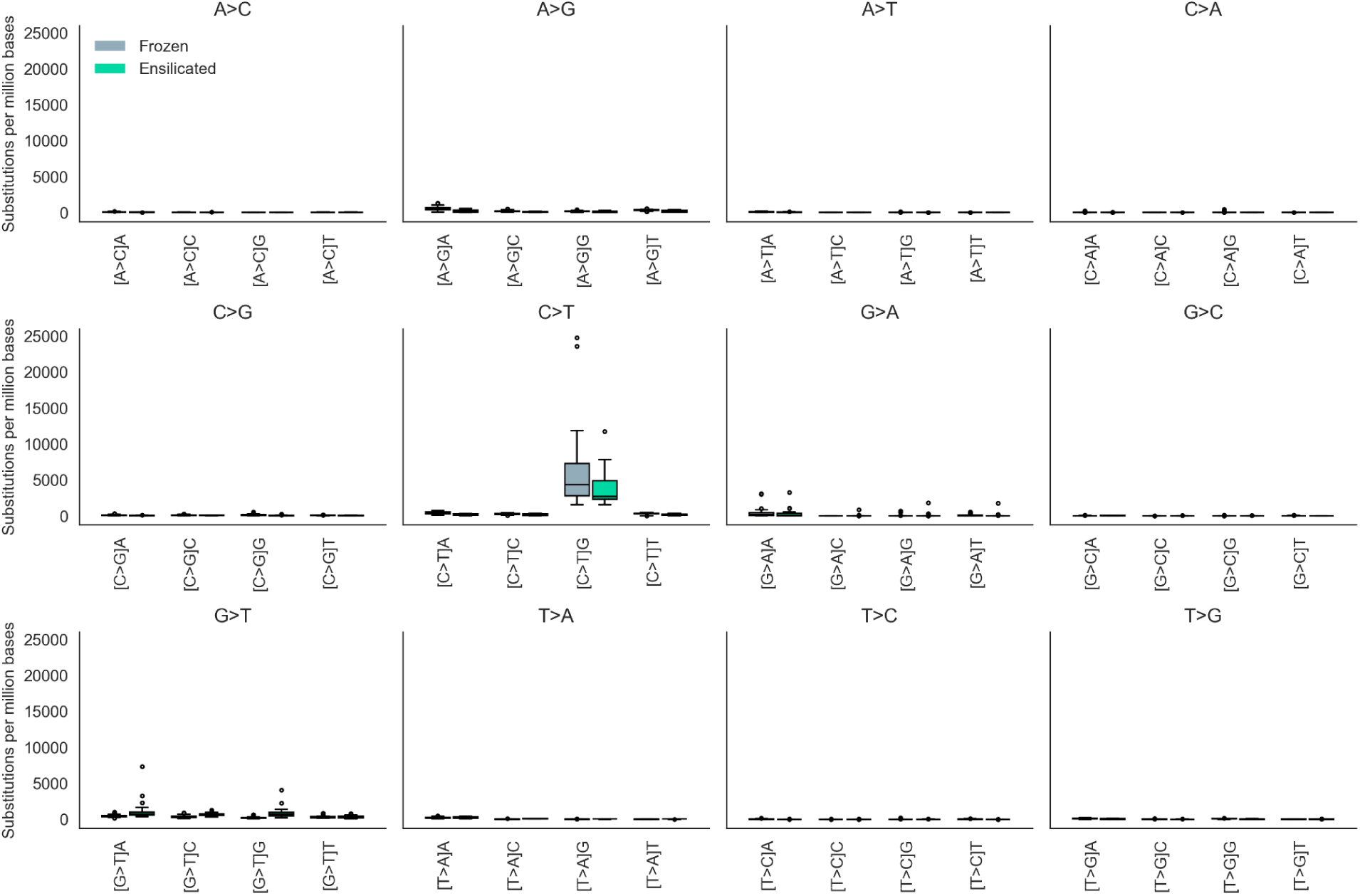
Substitution rates with post-dinucleotide context from tumor tissue.

**Supplementary Figure 5.**
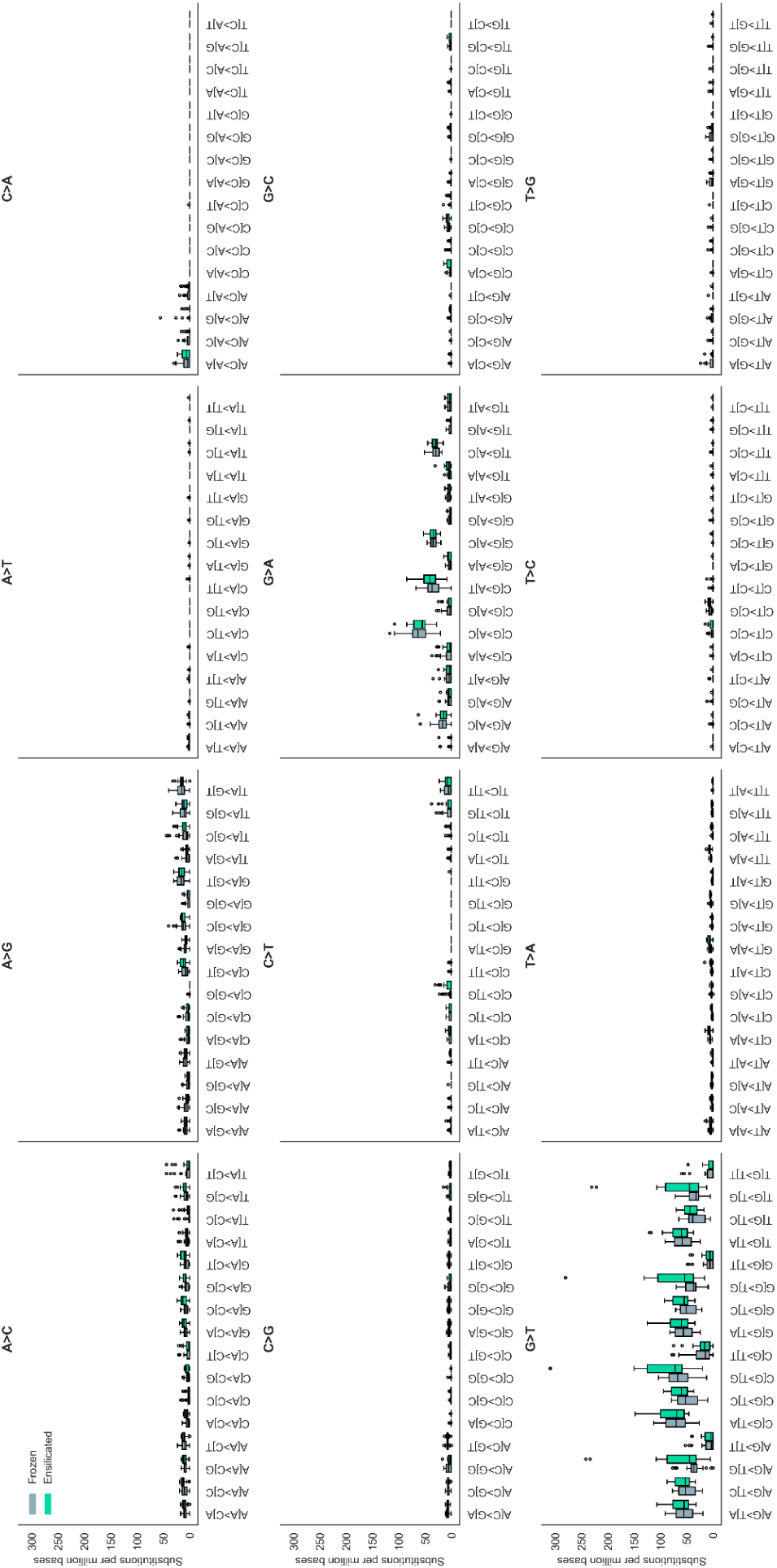
Substitution rates with trinucleotide context from normal tissue.

**Supplementary Figure 6.**
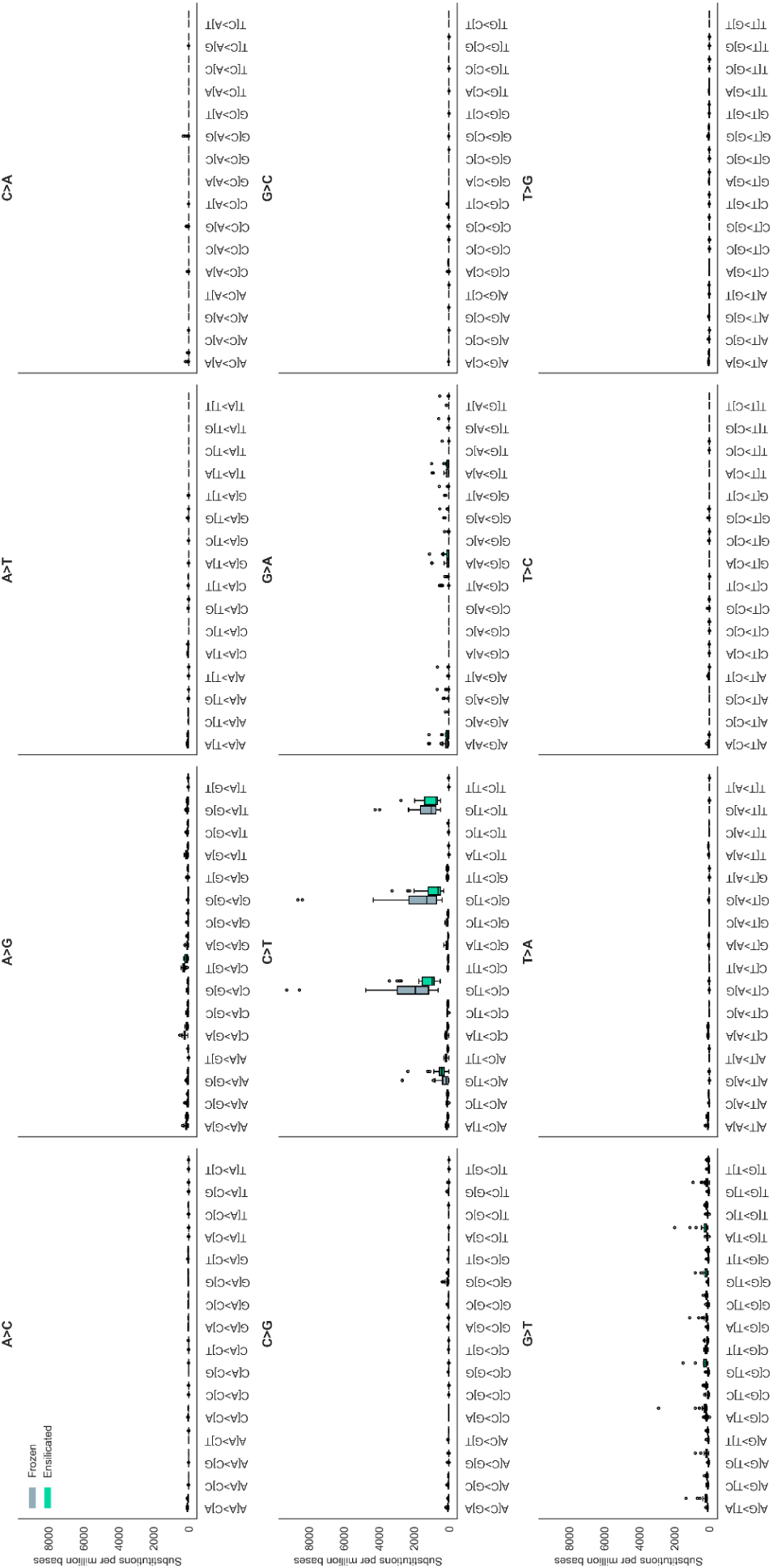
Substitution rates with trinucleotide context from tumor tissue.

**Supplementary Figure 7.**
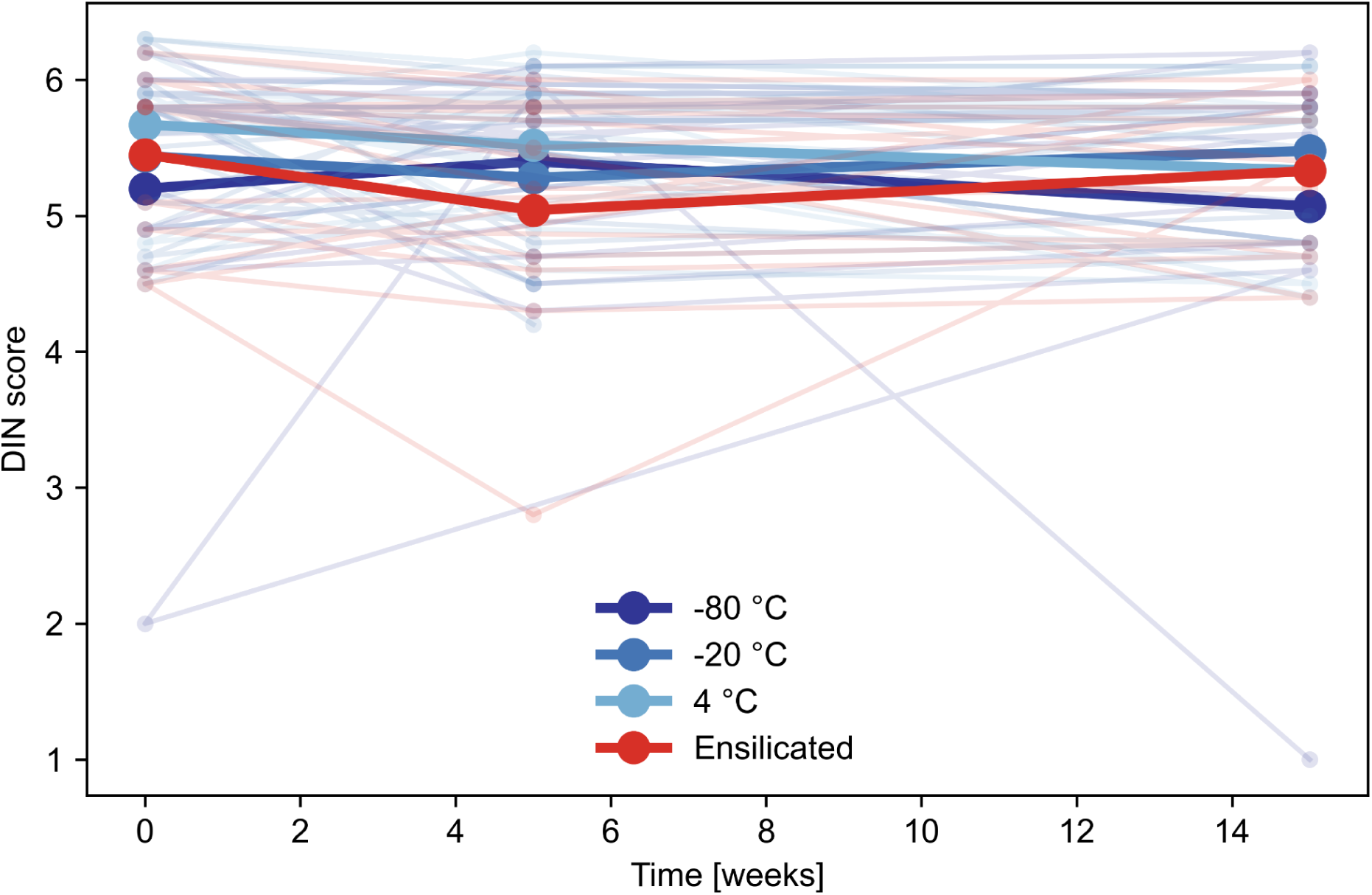
Ensilication preserves DNA integrity at room temperature comparable to frozen storage. Temporal profiles of DIN scores for DNA samples stored at -80 °C, -20 °C, 4°C, or ensilicated at room temperature over 15 weeks, with individual samples (thin lines) and means (thick lines) shown for each storage condition.

**Supplementary Table 1.** Library Insert sizes

| Sample | Metric | Normal frozen | Normal ensilicated | Tumor frozen | Tumor ensilicated |
| --- | --- | --- | --- | --- | --- |
| Replicate #1 | Median Insert Size | 222 | 211 | 146 | 152 |
|  | MAD | 60 | 74 | 33 | 40 |
| Replicate #2 | Median Insert Size | 227 | 226 | 137 | 160 |
|  | MAD | 63 | 69 | 36 | 42 |

**Supplementary Table 2.** Chimera rate

| Sample | Metric | Normal frozen | Normal ensilicated | Tumor frozen | Tumor ensilicated |
| --- | --- | --- | --- | --- | --- |
| Replicate #1 | Chimera Rate | 0.02941 | 0.029366 | 0.084617 | 0.042161 |
| Replicate #2 | Chimera Rate | 0.03311 | 0.029413 | 0.095364 | 0.060243 |

**Supplementary Table 3.** Sequencing library characteristics

| Sample | Metric | Normal frozen | Normal ensilicated | Tumor frozen | Tumor ensilicated |
| --- | --- | --- | --- | --- | --- |
| Replicate #1 | Library size | 178,749,102 | 247,144,894 | 77,892,967 | 58,233,756 |
|  | Duplication rate | 0.059442 | 0.062451 | 0.178443 | 0.213537 |
| Replicate #2 | Library size | 189,887,539 | 198,476,013 | 56,7816,85 | 35,038,012 |
|  | Duplication rate | 0.065969 | 0.071659 | 0.218177 | 0.301617 |

**Supplementary Table 4.** Mann-Whitney U statistic for normal samples

| Substitution | Frozen median | Ensiliated median | Median difference | U statistic | P-value |
| --- | --- | --- | --- | --- | --- |
| A>C | 80.5 | 136 | -55.5 | 295.5 | 0.01524760467 |
| A>G | 109.5 | 118 | -8.5 | 439.5 | 0.728865659 |
| A>T | 2 | 2 | 0 | 480.5 | 0.8133620896 |
| C>A | 4 | 7 | -3 | 457 | 0.9221326157 |
| C>G | 53.5 | 43 | 10.5 | 582 | 0.08954410397 |
| C>T | 15 | 17 | -2 | 446.5 | 0.8055086652 |
| G>A | 217.5 | 211 | 6.5 | 469 | 0.9481808024 |
| G>C | 9.5 | 15 | -5.5 | 381 | 0.2329523436 |
| G>T | 627 | 803 | -176 | 311 | 0.02763494753 |
| T>A | 35.5 | 52 | -16.5 | 192 | 8.71E-05 |
| T>C | 5 | 7 | -2 | 385 | 0.2558639309 |
| T>G | 10 | 3 | 7 | 696.5 | 0.00074025285 |

**Supplementary Table 5.** Mann-Whitney U statistic for tumor samples

| Substitution | Frozen median | Ensiliated median | Median difference | U statistic | P-value |
| --- | --- | --- | --- | --- | --- |
| A>C | 148.5 | 126 | 22.5 | 490 | 0.7073 |
| A>G | 1324 | 586 | 738 | 772 | 0 |
| A>T | 116.5 | 61 | 55.5 | 697 | 0.0008 |
| C>A | 0 | 0 | 0 | 527 | 0.1935 |
| C>G | 401 | 152 | 249 | 778 | 0 |
| C>T | 5466.5 | 3307 | 2159.5 | 656 | 0.0057 |
| G>A | 341 | 154 | 187 | 562 | 0.1561 |
| G>C | 25.5 | 56 | -30.5 | 304 | 0.0216 |
| G>T | 1260.5 | 2533 | -1272.5 | 115 | 0 |
| T>A | 263 | 308 | -45 | 368 | 0.1678 |
| T>C | 54.5 | 3 | 51.5 | 768 | 0 |
| T>G | 105 | 55 | 50 | 634 | 0.0147 |

**Supplementary Table 6.**
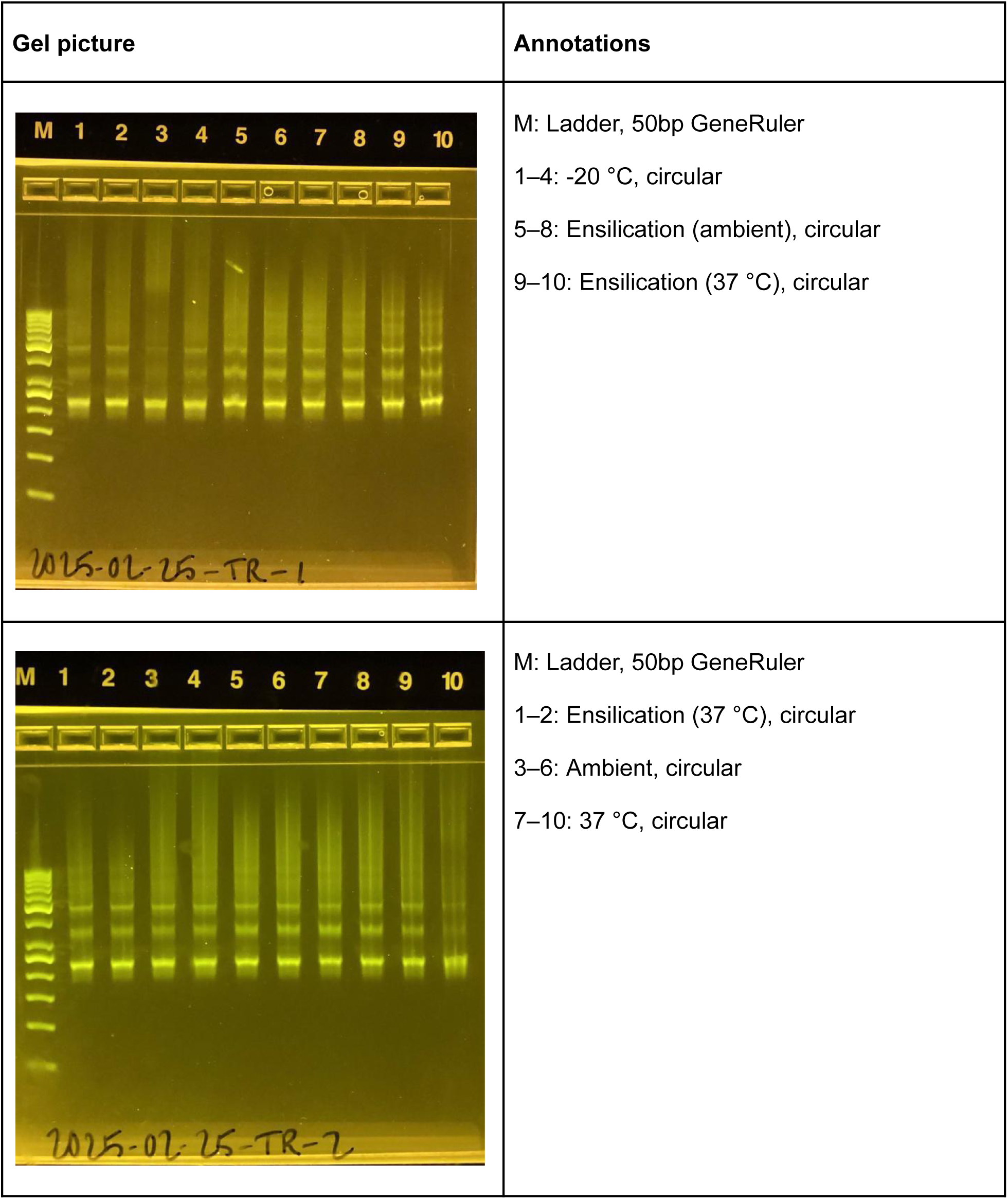
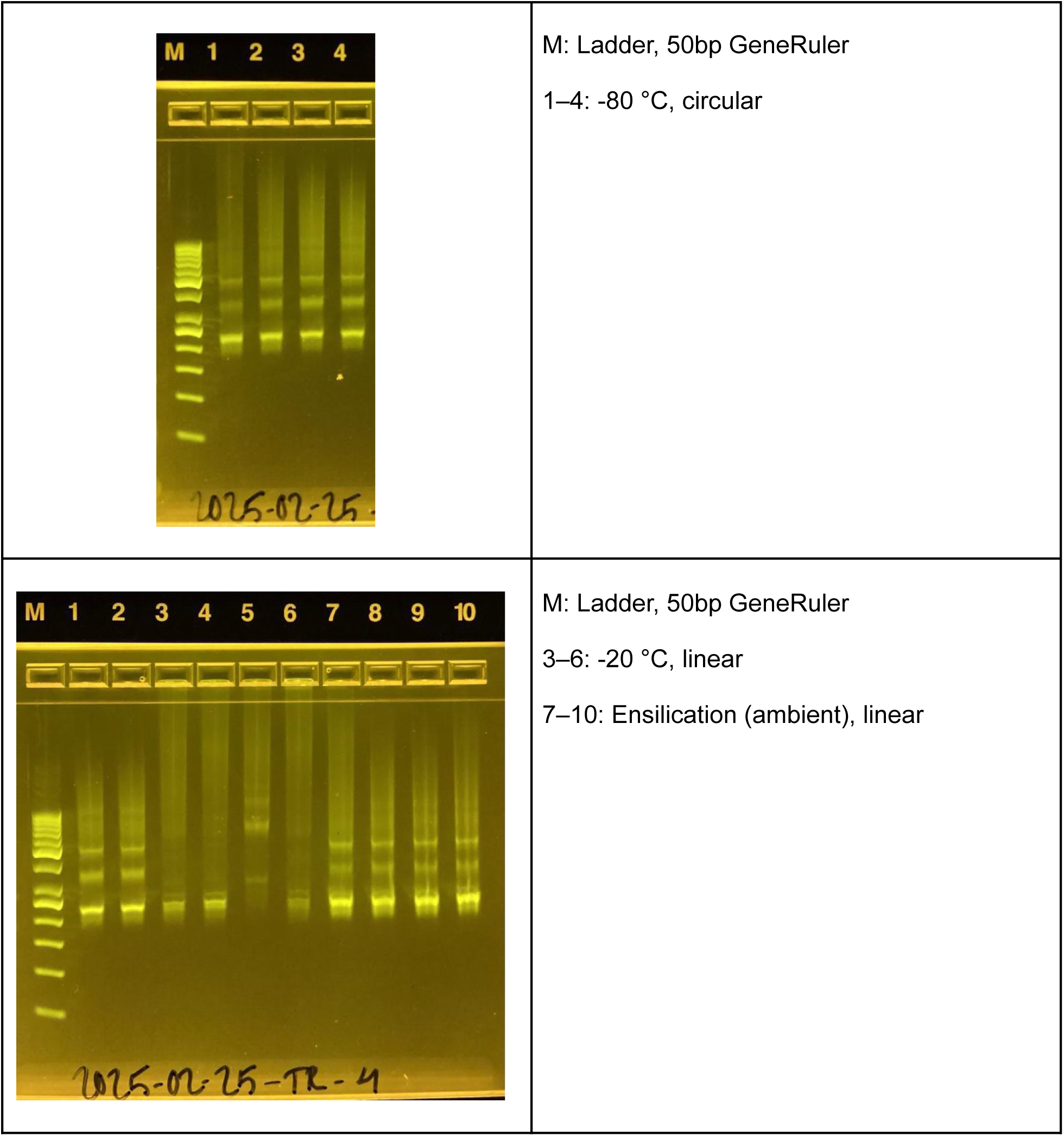
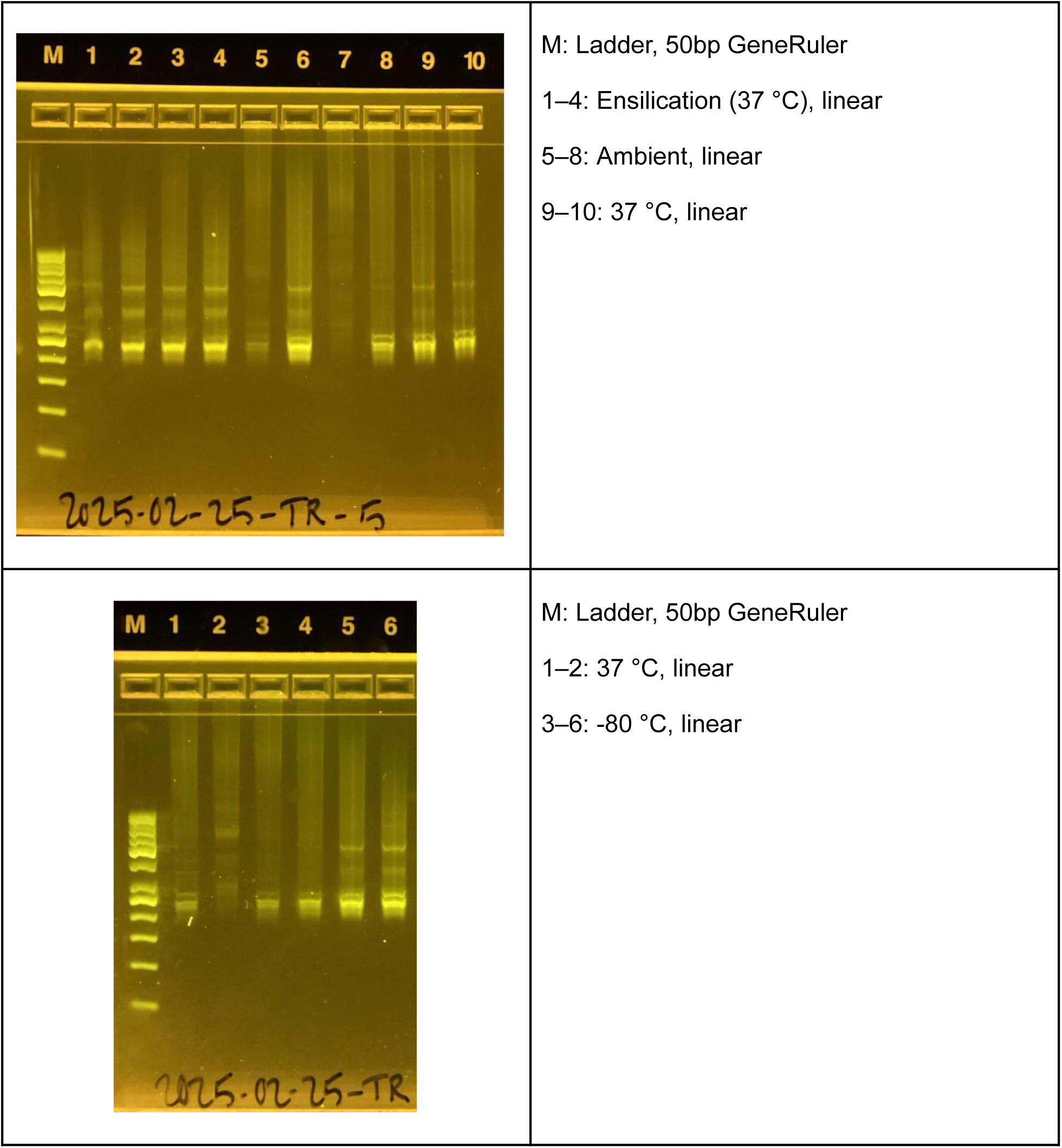
Preservation regardless of shape (circular ssDNA or linear). Successful preservation and sequencing of both linear and peanut-shaped circular ssDNA over time over many conditions showing ensilication is similar between -80 °C, -20 °C, ambient temperature, and 37 °C for 20 days. For linear DNA, ensilication far outperforms unprotected samples.

